# Characterization of Immune Responses to rAAVrh8 Gene Therapy for GM2 Gangliosidosis in Phase 1/2 Trial

**DOI:** 10.1101/2025.05.07.25327170

**Authors:** Terence R. Flotte, Meghan Blackwood, Motahareh Arjomandnejad, Ashley Harkins, Katelyn Sylvia, Sukanya Iyer, Danielle Kokoski, Rebecca Artinian, Allison M. Keeler

## Abstract

Understanding how the immune system responds to adreno-associated virus (AAV) gene therapy and potentially modulating that response is vital to their safety and ultimate success. However, the immune response in the central nervous system (CNS) to AAV gene therapy is still not well understood. Here, we characterized the immune responses to AAVrh8 vectors injected into the thalamus and cerebral spinal fluid (CSF) of Tay-Sachs (TSD) and Sandhoff (SD) disease patients. Nine patients in four dose cohorts were treated with gene therapy while being immunosuppressed with rituximab, sirolimus and prednisolone. Neutralizing antibodies against AAV capsid were detected in the serum of 9/9 patients and in the CSF of 7/9 patients. Specific T-cell responses against the AAV capsid were documented in all patients, with most patients developing responses at 2–3 weeks post-injection. Flow cytometry suggested the induction of capsid-specific regulatory T-cells in the periphery. Local immune responses were detected by cytokine analysis of the CSF along with upregulation of several chemokines, including CXCL8, CXCL9 and CXCL10. These Phase I/II clinical trial data provide valuable insights into how the human immune system responds to direct administration of AAV into the CNS and important assessments on the efficacy of the immune suppression regimen which can be used to inform future AAV clinical trials.

## Introduction

Characterization of immune responses to AAV gene therapy is critical for our understanding of how to design better gene therapies and their translation to clinical trials. Adeno-associated virus (AAV) gene therapy is a leading technology for the treatment of single gene disorders and has been associated with a relative safety profile. However, AAV is recognized by the immune system as a pathogen, and immunogenic responses have been associated with toxicities in some clinical trials[1, 2]. Many of these adverse immune events, such as the generation of capsid specific T-cells, were first observed in clinical trials because they had not been predicted by animal studies [3–7], thrombotic microangiopathy (TMA)[8, 9] and endothelial leakage[10]. Understanding how immune responses contribute to adverse events and changes in therapeutic protein expression over time is critical to refine vector designs and clinical trial protocols to increase safety and achieve durable clinical benefit in patients.

CNS-directed AAV clinical trials have not always robustly characterized immune responses[11]. AAV delivery to the CNS is considered a less immunogenic delivery route due to a unique immune environment in the brain such as lower Major Histocompatibility Complex (MHC) expression, expression anti-inflammatory factors and the blood-brain barrier (BBB)[11, 12]. However the CNS is not “immune privileged” and recent data have suggested that the there is a complex relationship between the immune system and the CNS[13]. The CNS has specialized immune cells, such as microglia being the tissue resident macrophages[14], as well as typical immune cells, such as T-cells, B-cells, NK cells and dendritic cells, albeit at very small numbers[15–18].

Here, we characterized the immune responses of infantile and juvenile patients receiving AAV gene therapy for the treatment of GM2 gangliosidosis Tay-Sachs (TSD) and Sandhoff (SD) diseases. Similar to our previously reported expanded access clinical trial[19], a 1:1 formulation of AAVrh8-*HEXA* and AAVrh8-*<u>HEXB</u>* monocistronic vectors was injected bilaterally into the thalamus and CNS in nine patients enrolled in four different dosing cohorts. Systemic and local immune responses were elevated over time despite a strong immunosuppressive regimen. Findings from this study can inform future clinical trial design to mitigate immune responses affecting the safety and efficacy of gene therapies.

## Results

### Clinical Trial Design

Nine patients with TSD or SD were treated as part of an open label, non-randomized two-stage, dose-escalation and safety study of a single of rAAVrh8-*HEXA*/*HEXB* (NCT04669535 and NCT06614569). Specific details are reported in a companion manuscript describing the clinical trial[20]. In brief, the AAV vector formulation was delivered bilaterally into each patient’s thalamus on day 1 followed by CSF delivery on day 2, with 75% of the CSF dose injected in the cisterna magna and 25% at the thoraco-lumbar region. The dose range was from 1.2E+13 vector genomes (vg) to 8.1E+13 vg for the bi-thalamic dose and from 1.42E+14 vg/patient to 3.6E+14 vg/patient for the total dose.

### Immunosuppression

To suppress B-cell responses, patients were immunosuppressed with an one infusion of the anti-CD20 monoclonal antibody rituximab at 375 mg/m^2^ approximately 2 weeks prior to AAV gene delivery. Flow cytometry confirmed B-cell depletion to less than 5% of total peripheral blood lymphocytes. To maintain total serum IgG at or above 500 mg/dl, intravenous immune globulin (IVIG) at 1000 mg/kg was given as needed post-vector delivery. To suppress T-cell responses, an initial infusion of 10 mg/kg methylprednisolone steroids followed by oral prednisolone was given at 1–2 mg/kg/day and continued until approximately 12 weeks post-infusion, followed by a 4-week taper where adjustments were made based on transaminases values. Oral sirolimus was also given at 1.5 mg/m^2^/day, titrated to a trough level between 7–12 ng/ml and was given through 24 weeks post-infusion, followed by 4-week taper. Trimethoprim-sulfamethoxazole and lansoprazole were additionally given to patients as prophylaxis for pneumocystis pneumonia and steroid-related peptic disease, respectively, while on sirolimus and steroids.

### Safety and Platelet Responses

AAV treatment was well-tolerated, only 15 adverse events (AEs) being determined to be possibly or definitely related to vector; the full description of AEs and severe AEs (SAEs) are reported in the companion manuscript[20]. No AEs were found to be related to innate immune responses, including thrombocytopenia or activation of complement. Although an initial reduction of platelets was noted after AAV delivery in all patients by day 2 or week 1 (**Figure 1a**), all patients remained at or above the normal range for total platelet numbers throughout the study (**Figure 1b**). No clinically significant bleeding episodes were observed.

**Figure 1.**
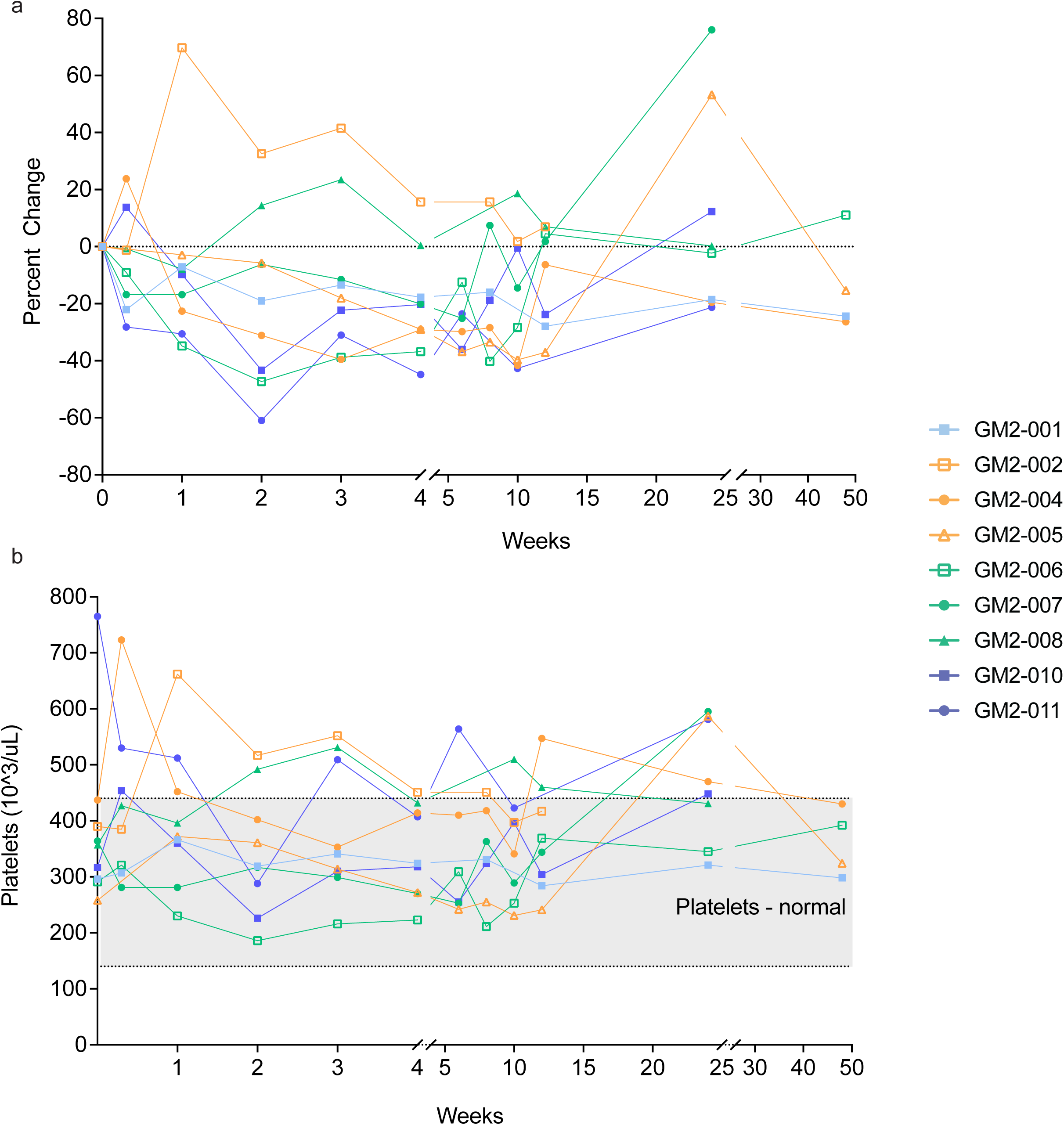
Percent Change and Total Platelet Count. Percent change in total platelet numbers in the blood was calculated from pre-dosing timepoint (**a**) Total platelet counts over time after AAV delivery (**b**). Gray shaded section designates normal range for platelet count. Colors designate dosage received: Light blue (starting dose) orange (low dose) green (mid dose) and dark blue (high dose). Open shapes indicate juvenile patients while closed shapes indicate infantile patients.

### Safety and Capsid Specific T-cell Responses

Infantile TSD patients have been reported to have elevated transaminase levels as part of the natural history of the disease[21], which was observed in our patients at baseline. Because at least two pre-treatment transaminase determinations per patient were available, we used these data points to create patient-specific baseline ranges. Adverse responses were defined as either elevations of at least 50% above the transaminase baseline values with a positive enzyme linked immunospot assay (ELISPOT) result or 100% above the transaminase baseline values without a positive ELISPOT result. Elevated liver function transaminases (LFTs) were noted after AAV administration in 0/1 patients at starting dose, 2/3 patients at low dose, 3/3 patients at mid dose and 2/2 patients at high dose. All patients developed capsid-specific interferon-gamma (IFNγ)-positive ELISPOTs in their peripheral blood mononuclear cells (PBMCs) during the course of the trial, with most patients testing positive at 2 weeks post-injection (range: 12–145 days) and often persisting throughout trial (**Figure 2a**).

**Figure 2.**
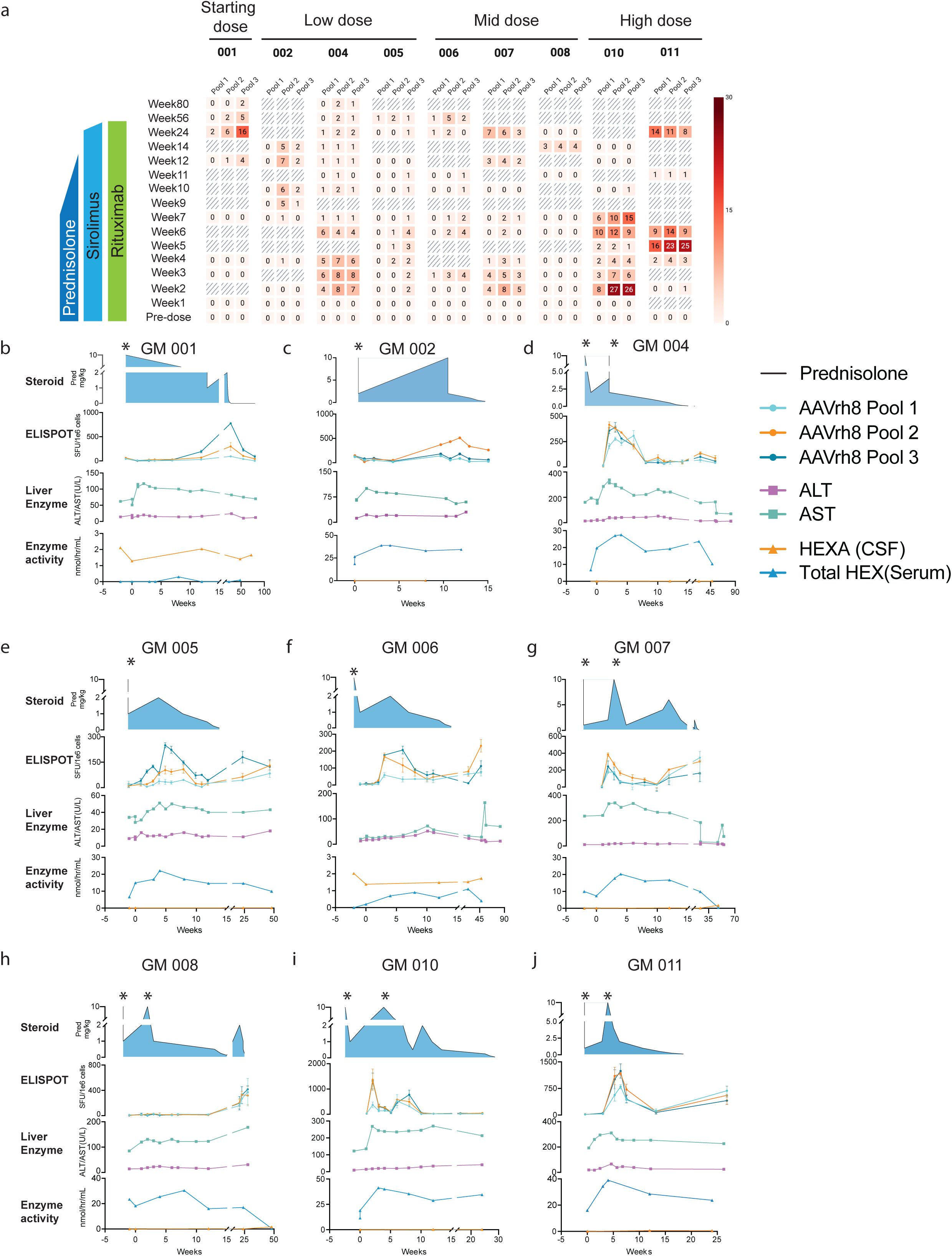
ELISPOTs Indicating Capsid Specific Immune Responses. Heat Map representation of IFNγ ELISPOT data (**a**). AAV rh8 capsid peptide are split between 3 pools. Gray diagonal lines designate timepoint where sample was not available. Positives are designated as 1 or higher with 1 standing for 3× the value of unstimulated control. Values are presented as the fold-increase over background. If 3× the unstimulated was less than 50 SFU /million cells than samples are considered positive or 1 at 50 SFU/million. General immune suppression regimen and timing is designated on the right. Individual patient (**b–j**) data graphed over time including steroid dosing, IFNγ ELISPOT SFU, ALT/AST Hexaminidase activity in the serum and HEXA activity in the CSF. *indicates 10 mg/kg methylprednisolone dosing; all other doses are oral prednisolone. SFU: spot forming units, ELISPOT: enzyme linked immunospot assay, ALT: alanine aminotransferase, AST: aspartate aminotransferase.

AAV capsid-specific IFNγ-positive ELISPOTs concomitant with elevated LFTs were observed in 1/2 patients at low dose, 2/3 patients at mid dose and 2/2 patients at high dose and were generally observed in the first 14–28 days post-AAV delivery. However, peak responses, as measured by spot forming units (SFU) on IFNγ ELISPOT, were observed between 2–28 weeks post-injection with the median peak at 4.9 weeks and secondary peaks observed in some patients between 23–47 weeks post-injection (**Figure 2b–j**). The patients receiving the highest dose had the most robust responses in SFUs (**Figure 2 a, i, j**) and the infantile patients (GM2-001,-004,-007,-008,-010,-011) had more robust responses compared to the juvenile patients (GM2-002,-005, 006) in general. Patients presenting with positive IFNγ ELISPOT responses and elevated liver enzymes were treated by increasing steroids either from 1 mg/kg to 2 mg/kg prednisolone (**Figure 2b,e,f**) or by a 3-day course of 10 mg/kg methylprednisolone (**Figure 2c,d,g,h,i,j**) followed by a return to 2 mg/kg dosage. ELISPOTs were reduced by steroid treatment, with reduced numbers of SFUs, but capsid-specific IFNγ producing cells were still detectable (**Figure 2**). Interestingly, during tapering of steroids, we frequently observed subsequent elevations of SFUs in most patients (**Figure 2b,c, e, f, g, h, j**). For the majority of patients, prednisolone tapering began at 12 weeks post-injection, but varied between 14 weeks post-injection up to 30 weeks post-injection. This was due to some patients requiring additional steroid dose increases and/or a prolonged taper of steroids (**Figure 2b–j**).

Two patients, GM2-006 and GM2-008, (**Figure 2f and h**) had elevated AST levels, elevated SFU numbers and decreased serum hexosaminidase (HEX) enzyme activity at timepoints after 24 weeks post-AAV delivery. In contrast only one patient, GM2-001, had an IFNγ immune response against the *HEXA* transgene (**Supplemental Figure 1**). Interestingly, this patient has SD with a mutation in *HEXB* but not *HEXA*. This IFN-γ-positive ELISPOT response against HEXA was transient and only observed at weeks 12 and 28. All ELISPOTs were performed with appropriate negative (unstimulated) and positive (CD3/CD28) controls to ensure the quality of T-cell responses (**Supplemental Figure 1**). Enzymatic activities of total HEX and HEXA were measured in the serum and CSF, respectively, for all patients over time (**Figure 2b–j**). Serum total HEX activity did not appear to be greatly affected by detection of capsid specific T-cells, particularly at early peaks of T-cell responses. HEXA activity in the CSF was very low in TSD patients (**Figure 2 c,d,e,g,h,i,j**). The HEXA assay detects the activity of both HEXαα and HEXαβ enzymes, and so the SD patients (GM-001 and GM-006) had higher enzymatic activity because they retain endogenous HEXαα activity (**Figure 2 b and f**).

### Capsid-Specific Antibody Responses

Neutralizing antibodies (Nabs) against rh8 capsid were measured in the serum of all patients prior to AAV delivery but were not used for exclusion criteria due to the route of injection being direct CNS delivery (**Figure 3**). Prior to AAV delivery, only patient GM2-002 had a positive titer at a dilution of 1:80. Despite peripheral B-cell depletion with rituximab being confirmed by clinical laboratory analysis, all patients seroconverted within 14 days of injection (**Figure 3a**). No differences in Nab titers were noted between doses or between juvenile or infantile patients. In general, Nab titers remained largely stable over time, but several patients had an upward trend after 24 weeks corresponding to the time when peripheral B-cells would be replenished after rituximab treatment. As for the additional immunosuppression, all patients stopped receiving steroids by 24 weeks post-injection, except for GM2-008 (stopped at 26 weeks) and GM2-010 (stopped at 30 weeks), however patients remained on sirolimus until 23–31 weeks post-injection.

**Figure 3.**
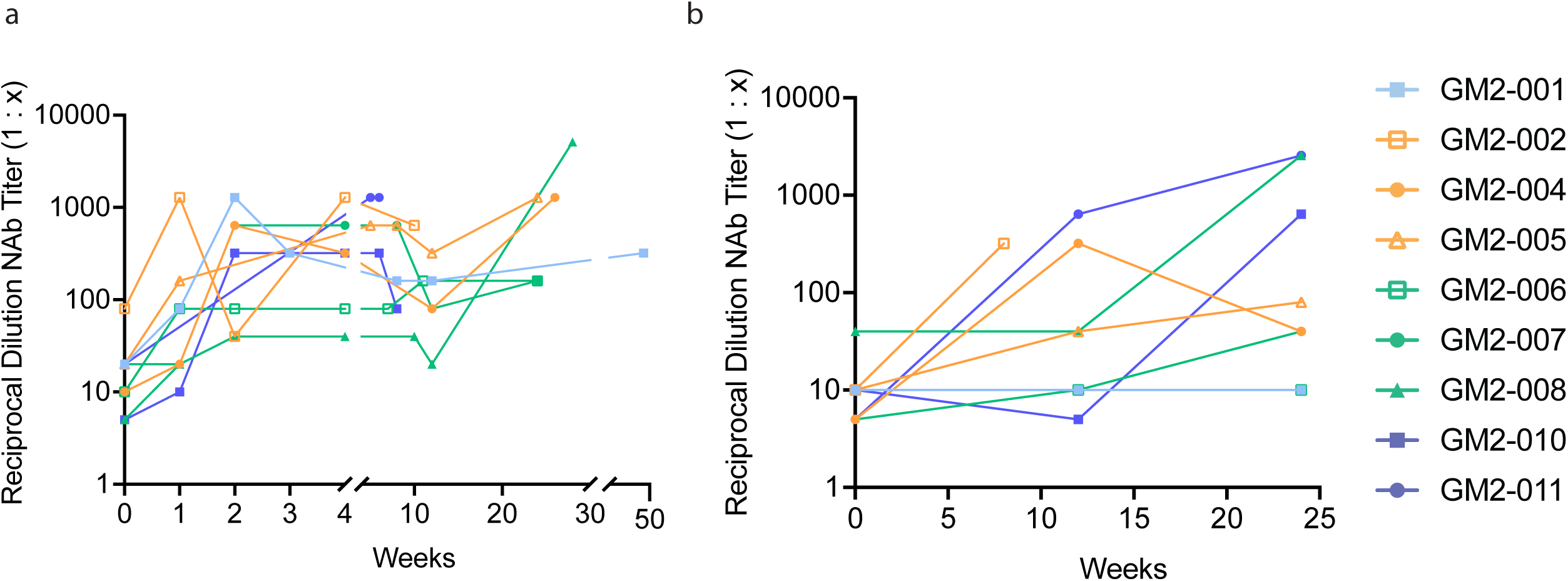
Neutralizing Antibody Titers in the Serum and CSF. Reciprocal neutralizing antibody (Nab) titers were measured in the serum (**a**) and CSF (**b**) for all patients over time. Colors designate dosage received: Light blue (starting dose) orange (low dose) green (mid dose) and dark blue (high dose). Open shapes are juvenile patients while closed shapes are infantile patients. All patients had titers over 1:20 (positive titer) in the serum (top) and 7/9 patients had positive titers in the CSF (bottom). Patients 001 and006 had titers of 1:10, which were considered a negative titer. Highest dilution measured for serum was serum was >1:5120 and for CSF was >1:1280, with some samples reaching upper limit of assay.

To further evaluate immune responses within the CNS, we evaluated Nab titers in the CSF (**Figure 3b**). Two patients, GM2-001 who received the low dose and GM2-006 who received the mid dose, remained seronegative with a titer of 1:10 in the CSF at all timepoints measured. The remaining patients all had positive titers in the CSF ranging from 1:40 to >1:1280. Because antibodies are not known to cross an intact BBB, this suggests that anti-capsid antibodies were generated from within the CNS. Total nucleated cells (TNC) and protein concentration were measured in the CSF of all patients. All patients had normal levels of both TNC and protein at all timepoints, except for GM2-005 at week 12 and GM2-011 at pre-screening. However, these samples contained high red blood cell counts (>100,000) suggesting blood contamination in the samples collected at those timepoints. Therefore, significant cellular infiltration into the CSF was not noted. Further imaging analyses also did not suggest cellular inflammation[20].

### Flow Cytometry

To further investigate the phenotype of capsid specific T-cells, flow cytometry analysis was performed to determine activation of different T-cell phenotypes after antigen stimulation. PBMCs were collected from two patients at two different timepoints, GM2-001 at 1 year and GM2-006 at week 4, and were cultured with either no stimulation, rh8 capsid peptide pools or positive stimulation of CD3/CD28 antibodies (**Figure 4a**). Flow cytometry was performed on the PBMCs to detect markers of activation and T-cell phenotype. A small induction of CD4^+^CD25^+^HELIOS+FoxP3^+^ population was observed after stimulation with the rh8 capsid peptide pools when compared to unstimulated controls (**Figure 4c**). Further, the percentage of activated CD25^+^ was increased in Tregs (defined as CD4+FOXP3+HELIOS+) after capsid stimulation but was not seen in CD4+ (defined as CD4^+^FOXP3^−^ HELIOS^−^) or CD8+ cells. However, all T-cells, Treg, CD4+ and CD8+ cells had upregulated expression of CD25 activation marker after CD3/CD28 stimulation, as a positive control (**Figure 4c**).

**Figure 4.**
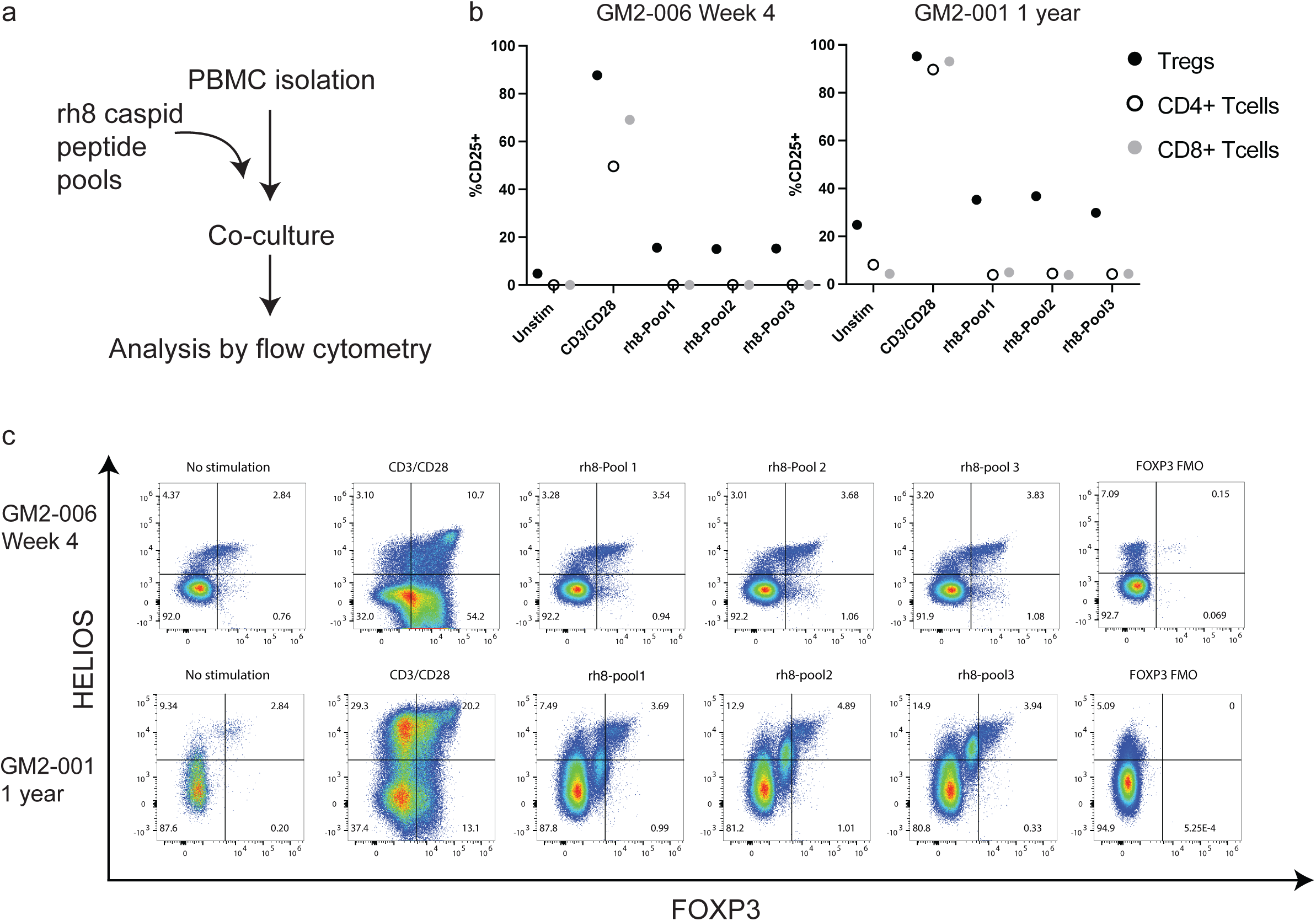
Capsid Specific Regulatory Cells in the Periphery. Patient PBMCs were stimulated with the same peptide pools used in ELISPOT experiments and then analyzed by flow cytometry (**a**). Quantification of cells positive for the CD25 activation marker (%CD25+) in two patients at two different timepoints (GM2-006 at week 4, left; GM2-001 at 1 year, right)(**b**). Backgating on all samples included singlets, live cells, lymphocyte gate and CD3+ cells. Tregs were further defined as CD4+FOXP3+HELIOS+, CD4+ T-cells were CD4+FOXP3-HELIOS- and CD8+ T-cells were gated on CD8+. Representative flow graphs from patients in (**b**) gated on live singlets, CD3+CD4+ showing increase of HELIOS+ FOXP3+ cells after stimulation with AAVrh8 peptide pools(**c**) Included no stimulation-negative control, CD3/CD28 positive control, three rh8 peptide pools and FOXP3 FMO control. PBMC: peripheral blood mononuclear cells, FMO-fluorescence minus one.

### Cytokine Evaluation

To further characterize immune responses in periphery and the CNS, cytokines and chemokines were evaluated in the serum (**Supplemental Figure 2**) and CSF (**Figure 5**, **Supplemental Figure 3**). Several chemokines were found to be elevated in the CSF of patients after AAV delivery, including CXCL8 and CXCL10 (**Figure 5**) by a multiplex flow-based bead assay. Elevations in chemokines in the CSF were further confirmed by an O-link assay, which suggested that CXCL9 and CXCL11 were also upregulated (**Supplemental Figure 3**). Cytokines IL-10 and IL-1β were elevated in the CSF, particularly at 24 weeks post-injection (**Figure 5**) by a multiplex flow-based bead assay. Additionally, IFNγ and PDL1 were elevated after AAV injection as determined by the O-link assay, whereas IL-6 was further found to be reduced after AAV delivery (**Supplemental Figure 3**).

**Figure 5:**
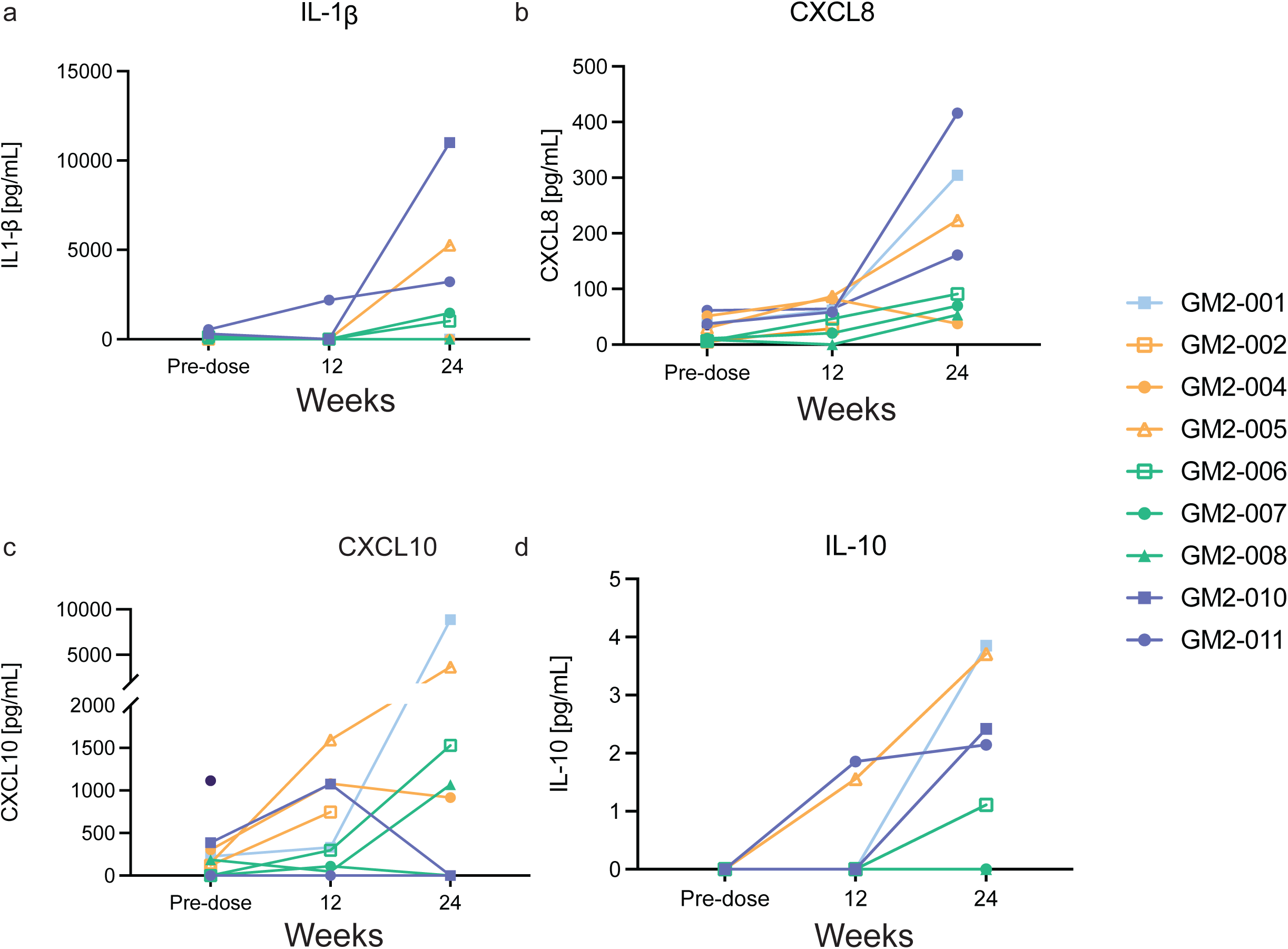
Chemokine and Cytokine evaluation of Cerebral Spinal Fluid. Cytokines and chemokines in the CSF were evaluated by multiplex cytometric bead array (CBA) method. CSF from four untreated GM2 gangliosidosis patients was pooled and run as disease state control. Colors designate dosage received: Light blue (starting dose) orange (low dose) green (mid dose) and dark blue (high dose). Open shapes indicate juvenile patients while closed shapes indicate infantile patients. Elevations were noted after gene therapy in 9/9 patients for CXCL9 (**a**), 8/9 patients for CXCL10(**b**) and 5/9 patients for both IL-10(**c**) and IL-1β (**d**).

## Discussion

Here, we describe TSD and SD patient immune responses to AAV gene therapy after a combined bi-thalamic and CSF delivery. Although this is a CNS-targeting route of injection, we believe that the vector leaked into the periphery because of the elevated total HEX activity, detection of Nabs and elevated ALT and AST in the serum.

Triple immunosuppression was utilized in this clinical trial, targeting both B- and T-cells. Despite rituximab eliminating circulating B-cells in the periphery, all patients developed Nabs in the serum within 2 weeks of AAV delivery. We note that our rituximab dosing, 375 mg/m^2^, was much lower than other rituximab dosing strategies, which delivered 1500 mg/m^2^ over multiple doses[22]. Nab titers did increase in several patients after 24 weeks when B-cells should be replenished in the periphery. Intriguingly, we observed a reduction of platelets from pre-treatment of between 20–60% in most patients after AAV delivery despite triple immunosuppression, yet the total platelet counts remained within the normal range for all patients at all timepoints.

All but two patients seroconverted to positive Nab titers in the CSF, suggesting cellular infiltration of B-cells to the CNS. By 12 and 24 weeks post-injection, 5/9 and 7/9 patients had positive Nabs in the CSF, respectively. All patients had increasing Nab titers between 12 and 24 weeks with the exception of the two patients who remained at a negative 1:10 titer. In a clinical trial to treat giant axonal neuropathy (GAN), AAV9 was administered by intrathecal injection at four different doses, which resulted in 13/14 patients having positive anti-AAV9 Nab titers and 13/14 patients having CSF white-cell pleocytosis, peaking by 3–6 months post-delivery[23]. Total dose ranges in the GAN trial were similar to the trial described here, between 3.5×10^13^–3.5×10^14^ vg, however the dosage in this TSD/SD trial was split between bi-thalamic and intrathecal routes. No evidence of white cell pleocytosis was observed in this TSD/SD clinical trial, as total nucleated cell counts and protein concentration were in the normal range, with two outliers showing evidence of contamination by contact with the blood during collection. Additionally in the GAN clinical trial, all patients received steroids, with some patients receiving rapamycin and patients that were cross-reactive immunologic material (CRIM)-negative receiving additional tacrolimus immunosuppression. The sub-clinical high elevation of white cell count in the CSF of one GAN patient was found to be steroid-responsive, and so an extended steroid regimen was utilized in all subsequent patients.

In the data reported here, we found that the capsid specific T-cell responses were steroid-responsive. Elevated T-cell responses observed in the first several weeks generally declined after increased steroid dosing. Other trials, such as the GAN study, have reported reduced capsid-specific T-cells with tacrolimus or combined tacrolimus/rapamycin immune suppression when compared to steroids alone[23]. Additionally in our study, several patients had capsid-specific T-cell responses at 24 weeks post-treatment and beyond which persisted or were elevated after the discontinuation of immunosuppression. Elevations in AST were also reported at these later timepoints, suggesting that capsid-specific T-cell responses persisted and potentially cleared transduced cells at several weeks post-delivery. Recently, transgene-directed immune-mediated myositis recurrence was observed after weaning patients off tacrolimus in a DMD clinical trial at approximately 1 year post-AAV delivery[24]. In response, the DMD patient was treated with an initial increased dosage of tacrolimus, but later was given intravenous methylprednisolone and rituximab due to worsening condition, and the patient remained on immunosuppression at the time of publication[24]. All patients in this GM2 trial were taken off immunosuppression and no clinical phenotype associated with immune responses to the gene therapy have been observed since.

Interestingly, Nab titers in either the CSF or serum did not suggest a dose response to vector. Clear dose responses were also not observed in the change in platelet numbers. However, the two patients in the highest dose generated the highest numbers of capsid-specific T-cells, suggesting a dose response. The CNS is a unique immune environment, and clinical trials have utilized this to their advantage[11]. Seropositive patients do not necessarily need to be excluded from direct CNS administration routes because circulating antibodies are unable to cross the blood brain barrier to prevent vector transduction. However, the brain is not as privileged from immune system complications as once believed and can mount robust immune responses. Robust T-cell responses to AAV capsid were observed in all patients at all doses despite the direct delivery route and immunosuppression regimen. While we were unable to assess the gene delivery to tissues in this study, our results strongly suggest that CNS-directed injection or infusion does not produce CNS-restricted biodistribution. The presence of elevated LFTs clearly is evidence of substantial vector biodistribution to the liver. Therefore, the lack of immune privilege may to some extent reflect the lack of true compartmentalization of vector delivery.

Most patients developed anti-capsid antibodies in the CSF after AAV administration, suggesting that B-cells in the CNS were producing antibodies because antibodies produced in the serum are unable to cross a functional BBB. Further, chemokine expression in the CSF suggests that some cellular infiltration may have occurred in the CNS. Cancer patients who receive Chimeric Antigen Receptor (CAR) T-cell therapies can experience immune effector cell-associated neurotoxicity syndrome, and in these cases patients can have significant neurotoxicity that is not observed in neuroimaging even at severe grades[25, 26]. However, evidence from our study suggests tolerance to the effector T-cell capsid-specific immune response. Induction of FoxP3+ cells were observed in patients at different timepoints after culturing T-cells with AAV capsid peptide pools. Induction of Tregs after delivery of AAV has been previously described after muscle delivery and is associated with stable transgene expression[27–30]. Therefore, we hypothesize that the CNS as a target for AAV gene therapy may be tolerogenic.

Several inferences may be drawn about the immune suppression management in this trial. First, the B-cell suppression used in this trial was not effective in preventing the development of NAb responses, however this may be correctable with higher doses of steroids, which has been reported by others to be more effective at reducing Nab responses. The second observation is that there are disease-specific considerations when using LFT values to indicate the need for increased steroids. In conditions with elevated baseline LFTs like infantile TSD, the availability of a sensitive and specific IFN γ ELISPOT assays are particularly important. Also, the quantification of ELISPOT responses (as opposed to positive/negative result reporting) allows for a more refined series of steroid dose adjustments.

Taken together, the data in this study gives us a better understanding of the way the that the immune system responds to a CNS-delivered gene therapy. Given the route of injection and immunosuppression, it was surprising that such robust capsid specific T-cells were observed in all patients. However, despite these responses the trial was safe and limited AEs were associated with AAV delivery. Further in-depth study and analysis will be necessary to better understand the role of the immune response on CNS directed gene therapies to produce better safety and efficacy profiles and achieve durable expression to correct disease pathologies.

## Methods

### Clinical Trial

The clinical trial design and protocol has been reported[20].

### Nabs

Serum and CSF were screened for neutralizing antibodies as previously described[19]. Luminescence was measured using a BioTek Synergy HTX reader. NAb titers are expressed as the highest dilution that inhibits β-galactosidase expression by at least 50% when compared to a negative mouse serum control.

### ELISPOT

IFNγ ELISPOT assays were performed as previously published[19]. The spot numbers were determined using a Mabtech IRIS reader. Responses were considered positive when the number of SFUs per 1×10^6^ cells were >50 and at least 3-fold higher than the control condition.

### Flow Cytometry

PBMCs were isolated from patients and rested for 24 hours. Following rest, cells were either left unstimulated or stimulated with rh8-pool1, rh8-pool2, rh8-pool3, HEX or anti-CD3/CD28 (Mabtech, Nacka Strand, Sweden) for 48 hours using a protocol similar to the ELISPOT assay. Single-cell suspensions were then stained for surface markers. Briefly, cells were suspended in 50 μL phosphate-buffered saline (PBS) with 1% fetal bovine serum (FBS) and incubated with human Fc block (BD Biosciences, Franklin Lakes, NJ) for 10 minutes at 4°C. After blocking, the appropriate antibodies for surface markers were added and incubated for 30 minutes at 4°C. Intracellular staining for CTLA-4, Helios and FOXP3 was performed using the FOXP3 staining kit (Invitrogen, Waltham, MA) according to the manufacturer’s instructions. Dead cells were excluded from all analyses using the Fixable Viability Kit. Data was acquired on an Aurora Cytek (Cytek Bio, Fremont, CA) and analyzed using FlowJo software v10.7.2. Antibodies used for flow cytometry are listed in Supplementary Table 1.

### CBA Assay

For the detection of cytokines in cerebrospinal fluid, a Cytometric Bead Array (CBA) assay was performed as previously described[10] using a Biolegend LEGENDplex HU Essential Immune Response Panel (13-plex) with V-bottom Plate (Biolegend, Cat# 740930) following the manufacturer’s recommendations. CSF samples from patients were not diluted for the assay. Data were acquired on a BD LSRII and analyzed using the LEGENDplex Data Analysis Software Suite.

## Data Availability

All data produced in the present study are available upon reasonable request to the authors.

## Acknowledgements

The authors are grateful to the patients and families who participated in this trial. In addition, we would like to thank Drs. Miguel Sena-Esteves, Heather Gray-Edwards, and Thomas Gallagher for their helpful comments on the manuscript.

## Contributions

AMK designed experiments, analyzed, interpreted data and wrote the paper. TRF, DK and RA were clinicians who were involved in clinical trial and provided samples, and TRF reviewed and edited the paper. MB, AH, KS and MA conducted experiments and analyzed data. SI helped with data visualization as well as presentation and reviewing the results.

## Ethics Statement

Ethical oversight of this study was provided by Western Institutional Review Board (Western IRB). The vector is being studied under Investigational New Drug (IND) #19314. All versions of the study as amended were approved by the Institutional Biosafety Committee of UMass Chan Medical School, WCG IRB, and Massachusetts General Hospital IRB. All patients were enrolled into this study after informed consent was obtained from their parents/guardians. These trials are registered on clinicaltrials.gov under identifiers NCT04669535 and NCT06614569.

---

**Supplemental Figure 1:**
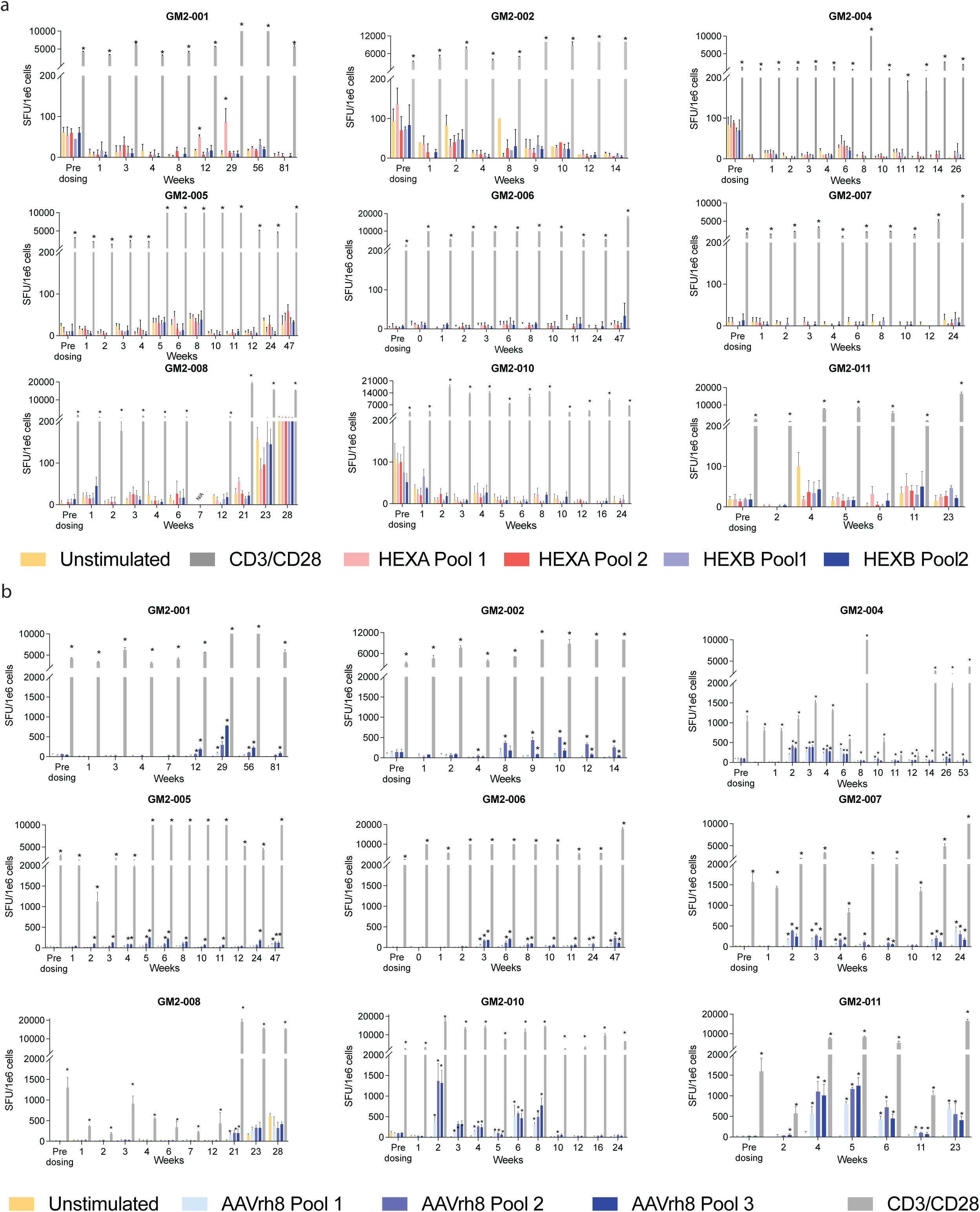
Transgene Specific and Capsid ELISPOT Response. Bar graph representation of individual IFNγ ELISPOT results from all timepoints taken from each patient. *indicates positive ELISPOT which is defined as 3× unstimulated control or 50 SFU/million cells if 3× unstimulated control is below 50 SFU/mL. CD3/CD28 antibody stimulation was used as positive control. Transgene or AAVrh8 capsid peptide pools were generated with 15 amino acids per peptide with a five amino acid offset to cover entire sequence of delivered HEXA and HEXB cargo or entire AAVrh8 capsid sequence. Peptides were split between three pools with no more than 50 peptides per pool. SFU: spot forming unit, ELISPOT: enzyme linked immunospot assay.

**Supplemental Figure 2.**
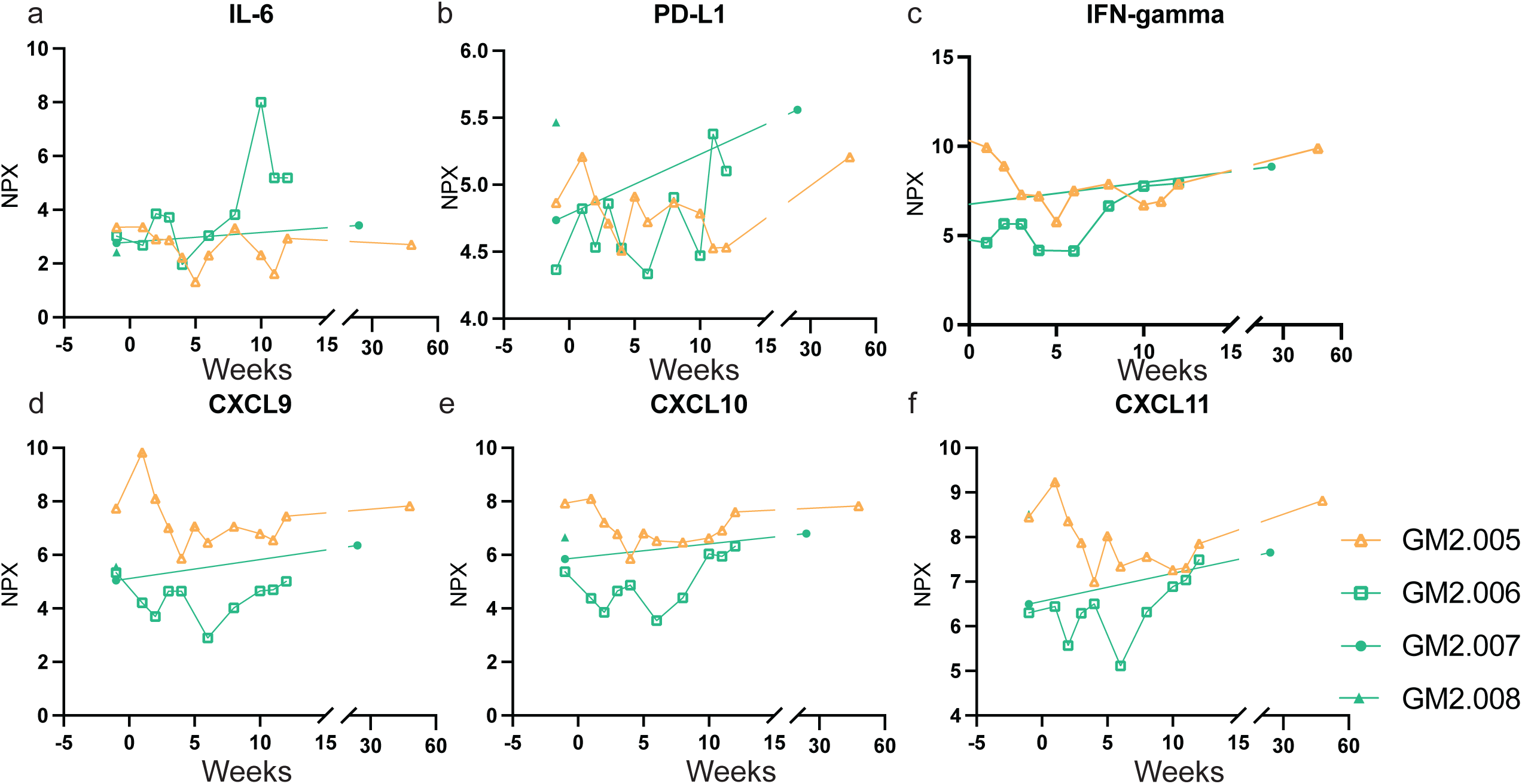
Serum Chemokine and Cytokine O-link Expression Data. Serum chemokines and cytokines were evaluated using an O-link 96 inflammation panel. Samples from four patients were analyzed by this assay along with untreated GM2 disease controls. Normalized protein expression (NPX) is shown at log_2_ scale.

**Supplemental Figure 3.**
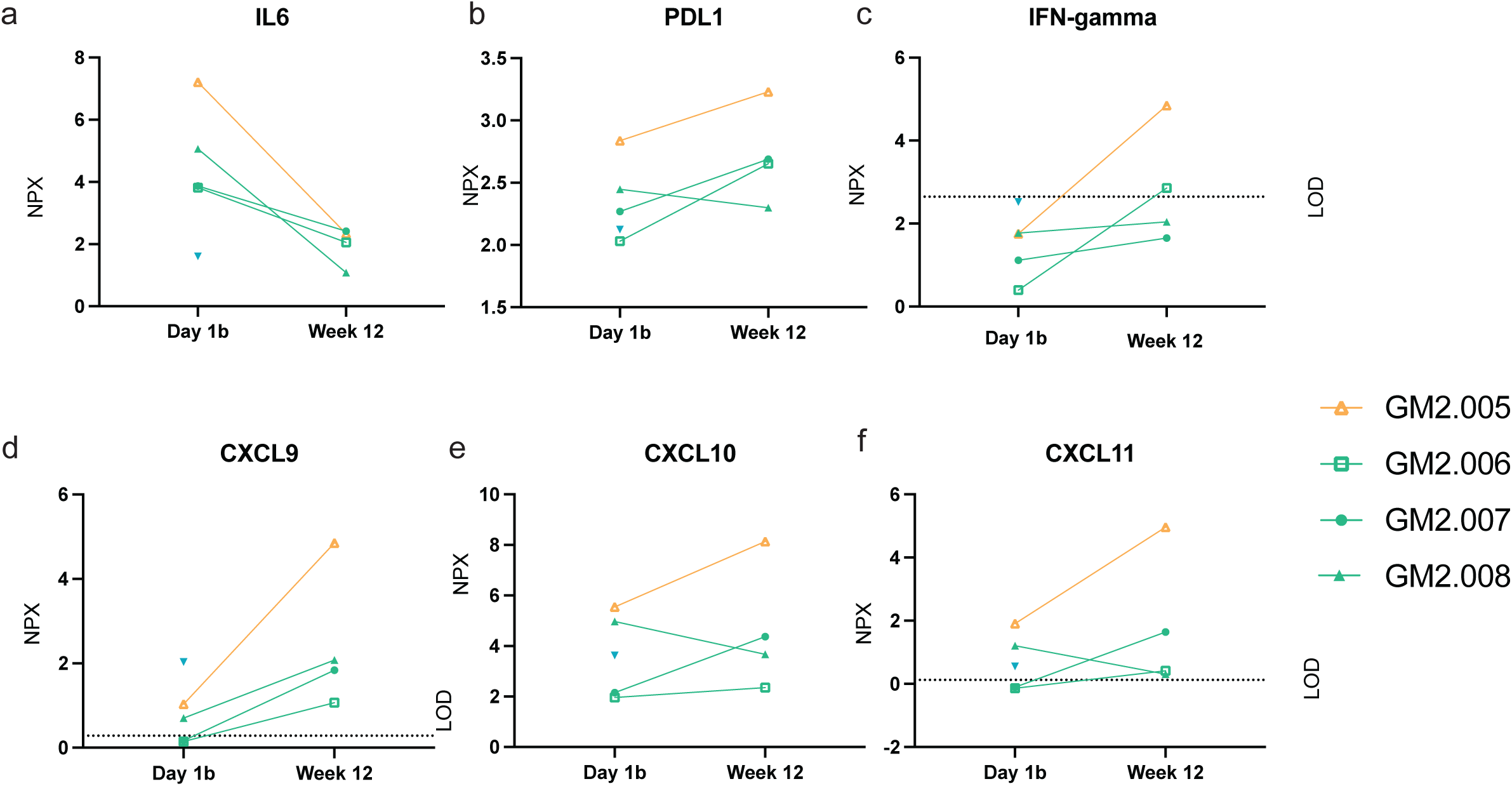
CSF Chemokine and Cytokine O-link Expression Data. CSF chemokines and cytokines were evaluated using an O-link 96 inflammation panel. Samples from four patients at two timepoints were analyzed by this assay to validate the CBA results along with GM2 disease controls. Normalized protein expression (NPX) is shown at log_2_ scale. Limit of detection (LOD) for this assay is indicated as designated by the manufacturer, and some values reported were below this level.

